# Glucose metabolism reflects local atrophy and tau pathology in symptomatic Alzheimer’s disease

**DOI:** 10.1101/2021.03.08.21252999

**Authors:** Amelia Strom, Leonardo Iaccarino, Lauren Edwards, Orit H. Lesman-Segev, David N. Soleimani-Meigooni, Julie Pham, Suzanne L. Baker, Susan Landau, William J. Jagust, Bruce L. Miller, Howard J. Rosen, Maria Luisa Gorno-Tempini, Gil D. Rabinovici, Renaud La Joie, for the Alzheimer’s Disease Neuroimaging Initiative

## Abstract

Posterior cortical hypometabolism measured with [^18^F]-Fluorodeoxyglucose (FDG)-PET is a well-known marker of Alzheimer’s disease-associated neurodegeneration, but its associations with underlying neuropathological processes are unclear. We assessed the relative contributions of three potential mechanisms causing hypometabolism in the retrosplenial and inferior parietal cortices: local molecular (amyloid and tau) pathology and atrophy, distant factors including contributions from the degenerating medial temporal lobe or molecular pathology in functionally connected regions, and the presence of the apolipoprotein E (*APOE)* ε4 allele. Two hundred and thirty-two amyloid-positive cognitively impaired patients from two independent cohorts (University of California, San Francisco, UCSF, and Alzheimer’s Disease Neuroimaging Initiative, ADNI) underwent MRI and PET with FDG, amyloid-PET using [^11^C]-Pittsburgh Compound B, [^18^F]-Florbetapir, or [^18^F]-Florbetaben, and [^18^F]-Flortaucipir tau-PET within one year. Standard uptake value ratios (SUVR) were calculated using tracer-specific reference regions. Brain regions were defined in native space using FreeSurfer. Regression analyses were run within cohorts to identify variables associated with retrosplenial or inferior parietal FDG SUVR. On average, ADNI patients were older and had less severe cognitive impairment than UCSF patients. Regional patterns of hypometabolism were similar between cohorts, though there were cohort differences in regional gray matter atrophy. Local cortical thickness and tau-PET (but not amyloid-PET) were independently associated with both retrosplenial and inferior parietal FDG SUVR (Δ*R*^*2*^ = .09 to .21) across cohorts in models that also included age and disease severity (local model). Including medial temporal lobe volume improved the retrosplenial FDG model in ADNI (Δ*R*^*2*^ = .04, *p* = .008) but not UCSF (Δ*R*^*2*^ < .01, *p* = .52), and did not improve the inferior parietal models (Δ*R*^*2*^s < .01, *p*s > .37). Interaction analyses revealed that medial temporal volume was more strongly associated with retrosplenial FDG SUVR at earlier disease stages (*p* = .06 in UCSF, *p* = .046 in ADNI). Models including molecular pathology in functionally connected regions, defined based on task-free functional MRI data from the Neurosynth database, or *APOE* ε4 did not outperform local models. Overall, hypometabolism in cognitively impaired patients primarily reflected local atrophy and tau pathology, with an added contribution of medial temporal lobe degeneration at earlier disease stages. Our data did not support hypotheses of a detrimental effect of pathology in connected regions or the presence of the *APOE* ε4 allele in impaired participants. FDG-PET reflects structural neurodegeneration and tau, but not amyloid, pathology at symptomatic stages of Alzheimer’s disease.

## Introduction

Alzheimer’s disease is defined as the pathological accumulation of β-amyloid plaques and neurofibrillary tau tangles in the brain, which are thought to induce neurodegeneration and cognitive decline.^1^ FDG-PET is a marker of glucose metabolism and is believed to largely reflect neuronal and synaptic activity.^2^ Patients with Alzheimer’s disease frequently exhibit a pattern of temporo-parietal hypometabolism on FDG-PET indicating neurodegeneration.^3–7^ Regional patterns of decreased FDG-PET correlate with cognitive and functional impairment^8–12^ and are detectable early in the disease course, even before clinical symptoms.^13,14^ There are multiple mechanisms, perhaps coexisting, that could cause brain glucose hypometabolism (Figure 1A).

**Figure 1.**
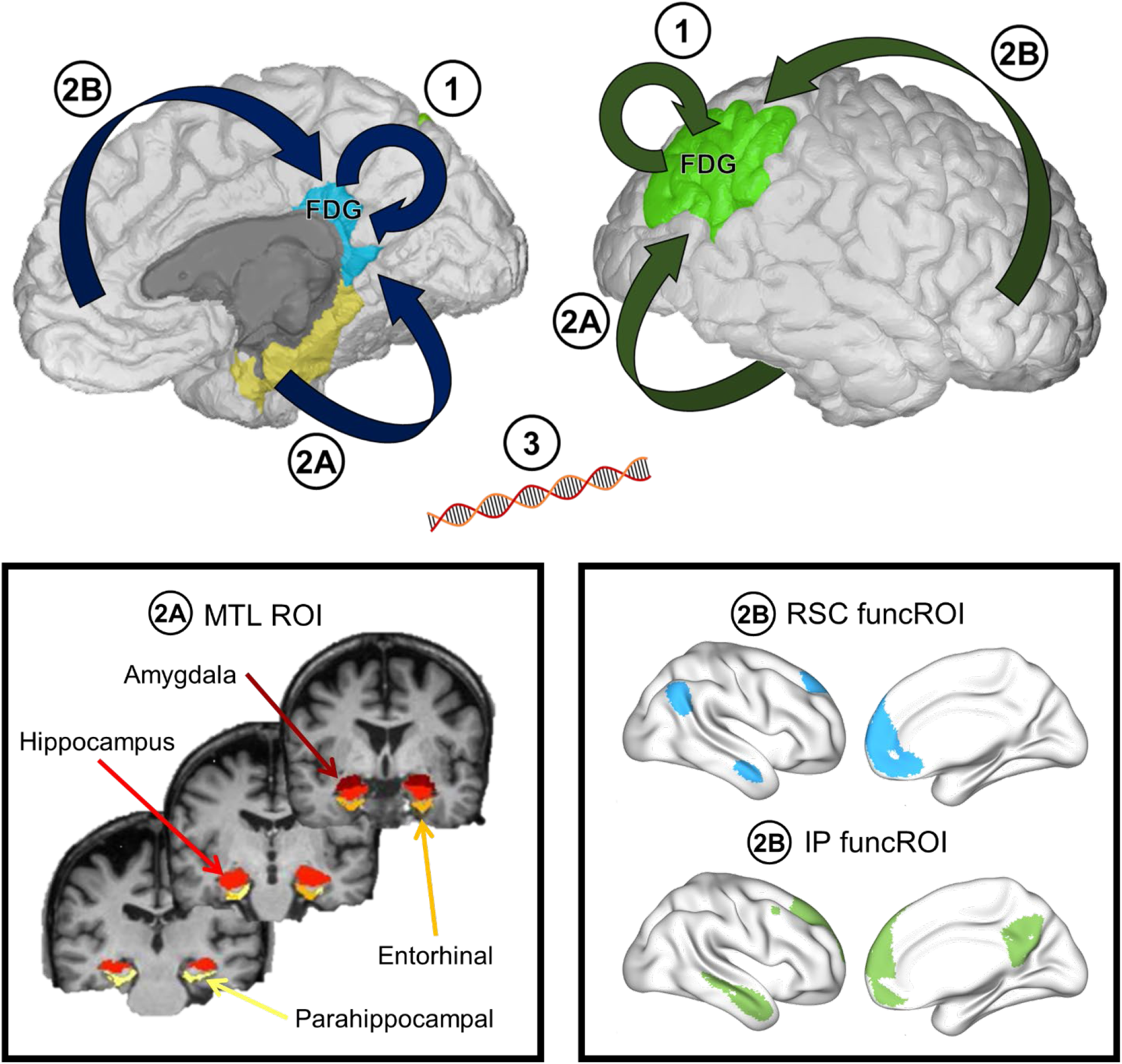
Schematic of potential hypometabolism-related mechanisms and regions of interest used. Hypometabolism in the primary regions of interest (blue = retrosplenial cortex, green = inferior parietal lobe) may result from local molecular pathology and atrophy (1), from distant effects of medial temporal atrophy (2A) or molecular pathology in functionally connected regions (2B), and/or from the presence of the *APOE* ε4 allele (3). Yellow on top panel = medial temporal lobe. The medial temporal lobe ROI consists of the amygdala, hippocampus, entorhinal cortex, and parahippocampal cortex (bottom left panel). The funcROIs were derived from resting-state functional data from healthy controls for each primary region of interest separately (bottom right panel).

First, molecular pathology (amyloid and tau) and degeneration are associated with hypometabolism within a region (local). Lower brain volume consistently correlates locally with decreased FDG-PET.^5,15–18^ This association may be partially explained by increased partial volume effects, where the limited spatial resolution of PET underestimates local radiotracer retention especially in the setting of severe atrophy,^19,20^ though partial volume correction does not remove this association.^15^ However, regional hypometabolism in Alzheimer’s disease exceeds what is expected from, and does not perfectly overlap with, atrophy.^15,21,22^ Local tau pathology may partially explain this discrepancy in pattern because tau-PET is related to both structural MRI and FDG-PET measures, though more so to FDG-PET.^18,23,24^ Tau-PET patterns consistently overlap with decreased FDG-PET in preclinical and clinical populations.^4,22,25–27^ Some studies have demonstrated a local correlation between amyloid-PET and FDG-PET as well,^28–30^ but others have found no such association.^5,31–35^

Second, regional hypometabolism can be related to abnormalities occurring in distant regions. For example, FDG-PET can be decreased in regions that are physically distant from but downstream of a structural or pathological lesion.^36,37^ A lesion to the rhinal cortex in primates results in reduced glucose metabolism in many remote brain regions, including parietal, frontal, occipital, and cingulate cortices.^38^ Similarly, in human imaging studies of patients with Alzheimer’s dementia, MTL atrophy is related to reduced FDG-PET in the retrosplenial cortex (RSC), perhaps mediated by the disruption of the cingulum bundle.^28,35,39^ Molecular pathology may also influence neurodegeneration remotely via network connections, with supporting evidence for this phenomenon having been observed with tau- and amyloid-PET.^24,40,41^ Notably, Pascoal *et al*.^34^ found that hypometabolism was not related to local amyloid-PET signal, but was associated with amyloid-PET uptake in functionally connected regions.

Third, the *APOE* ε4 allele may itself be associated with reduced metabolism in Alzheimer’s disease regions, even in young adults or older adults without amyloid pathology, perhaps as an endophenotype.^42–46^ In symptomatic stages of Alzheimer’s disease, findings are more conflicting with some studies suggesting that *APOE* ε4 is associated with a greater decrease in metabolism in Alzheimer’s disease regions,^47,48^ while others find no such association.^30,31,49^

Previous findings about determinants of Alzheimer’s disease-related hypometabolism may be discrepant due to small samples, the inclusion of amyloid-positive and amyloid-negative patients, or the inclusion of both impaired and unimpaired participants. The present study investigates these potential determinants in two independent samples of amyloid-positive, clinically impaired patients (total *N* = 232) to understand their relative contributions to hypometabolism in Alzheimer’s disease. An understanding of these relationships is important to improve the interpretation of FDG-PET in both research studies and clinical contexts, where FDG-PET is commonly used.

We focused on three hypotheses formulated based on available literature: 1) local contributions from atrophy and molecular pathology, 2) distant effects from either the degenerating MTL (2a) or molecular pathology in functionally connected regions (2b), and 3) the presence of the *APOE* ε4 allele (Figure 1). To test these hypotheses, we focused on two regions that display consistently salient hypometabolism on FDG-PET in Alzheimer’s disease patients: the posterior cingulate / RSC and inferior parietal (IP) lobe.^3,6,7,50^ We performed analyses in amyloid-positive, cognitively impaired patients with Alzheimer’s disease from two complementary cohorts covering a range of ages, clinical criteria, and disease severity to strengthen the generalizability of our findings and to potentially understand some of the conflicting results in the existing literature.

We hypothesized that local atrophy, local tau pathology, MTL volume, and molecular pathology in connected regions would be associated with Alzheimer’s disease hypometabolism, as these relationships have been observed at symptomatic stages. We expected that MTL volume would be specifically associated with decreased FDG-PET in the RSC but not IP due to its robust structural connections with the RSC.^51^ However, we did not expect *APOE* ε4 to play a role, as its effect has most consistently been observed in preclinical stages.^45^

## Materials and methods

### Participants

Two hundred and thirty-two cognitively impaired individuals were retrospectively included from two independent cohorts. Selection criteria included i) a diagnosis of Mild Cognitive Impairment or dementia due to Alzheimer’s disease based on clinical information,^52,53^ ii) available MRI and PET with FDG, an amyloid tracer, and [^18^F]-Flortaucipir (FTP) within 1 year, and iii) amyloid-PET positivity.

The first cohort consisted of 85 patients from the UCSF Alzheimer’s Disease Research Center. Exclusion criteria for UCSF patients included a history of repetitive head trauma consistent with possible traumatic encephalopathy syndrome.^54^ Twenty-nine patients met additional criteria for specific variants of Alzheimer’s disease: logopenic variant of primary progressive aphasia (n=12) and posterior cortical atrophy (n=17). Amyloid-PET positivity was determined by both visual read by an expert neurologist and an SUVR quantitative threshold (composite score >1.21) of PET with [^11^C]-Pittsburgh Compound B (PIB).^55^

The second cohort consisted of 147 patients from the ADNI study (adni.loni.usc.edu). We included all available ADNI cases with a clinical diagnosis of Mild Cognitive Impairment (early or late) or Alzheimer’s disease (dementia) within 1 year of the imaging studies. Amyloid-PET positivity was determined via quantification using tracer-specific quantitative thresholds (see www.adni-info.org). The ADNI was launched in 2003 as a public-private partnership, led by Principal Investigator Michael W. Weiner, MD. The primary goal of ADNI has been to test whether serial MRI, PET, other biological markers, and clinical and neuropsychological assessment can be combined to measure the progression of Mild Cognitive Impairment and early Alzheimer’s disease. For up-to-date information, see www.adni-info.org.

Ethical approval for the present study was given by the University of California (San Francisco and Berkeley) and Lawrence Berkeley National Laboratory institutional review boards for human research, and written informed consent was obtained from all participants according to the Declaration of Helsinki.

### Image acquisition and pre-processing

#### UCSF

T1-weighted magnetization prepared rapid gradient echo MRI sequences were acquired for UCSF patients on either a 3T Siemens Tim Trio (n=30) or a 3T Siemens Prisma Fit scanner (n=55). Acquisition parameters were similar for both scanners (sagittal slice orientation; slice thickness = 1.0 mm; slices per slab = 160; in-plane resolution = 1.0×1.0 mm; matrix = 240×256; repetition time = 2,300 ms; inversion time = 900 ms; flip angle = 9°), although echo time slightly differed (Trio: 2.98 ms; Prisma: 2.9ms). Each participant’s MRI was segmented and parcellated using Freesurfer 5.3 to define ROIs and extract gray matter volume and cortical thickness measures.

All PET scans were acquired at the Lawrence Berkeley National Laboratory on a Siemens Biograph 6 Truepoint PET/CT scanner. PIB and FTP were synthesized and radiolabeled at the Laboratory’s Biomedical Isotope Facility. For FDG-PET scans, participants fasted for at least 6 hours before scanning. A low-dose CT scan was performed for attenuation correction, and PET data were reconstructed using an ordered subset expectation maximization algorithm with weighted attenuation with scatter correction and a 4 mm Gaussian kernel. PET SUVR images were based on mean uptake over tracer-specific acquisition windows post-injection (30-60 min for FDG, 50-70 min for PIB, and 80-100 min for FTP) normalized by mean uptake in MRI-defined (Freesurfer- and SUIT-based), tracer-specific reference regions (pons for FDG, cerebellar cortex for PIB, and inferior cerebellar cortex for FTP). Reconstructed image resolution was 6.5 x 6.5 x

7.25 mm. An additional 4 mm isotropic Gaussian filter was applied to smooth PET images to a final effective ∼8 mm^3^ resolution (matching ADNI scans, see below).

#### ADNI

MRI and PET acquisition protocols are detailed elsewhere, see www.adni-info.org. For the present study, we used MRI T1-weighted sequences that were segmented and parcellated by Freesurfer 5.3.

PET scans were acquired according to published protocols and analyzed using tracer-specific acquisition windows: 30-60 min for FDG, 50-70 min for [^18^F]-Florbetapir (FBP), 90-110 min for [^18^F]-Florbetaben (FBB), and 75-105 min for FTP. Reference regions used mirrored those of UCSF (pons for FDG, cerebellar gray for FBP and FBB, inferior cerebellar gray for FTP). We converted template-based FDG SUVR images from the ADNI database into MRI-based FDG SUVR images with a pons reference region defined via a custom pipeline (Supplementary Methods).

All image processing beyond Freesurfer parcellation, including PET pre-processing, warping, and MRI tissue probability segmentation, was performed using Statistical Parametric Mapping (SPM12; Wellcome Trust Center for Neuroimaging, London, UK, http://www.fil.ion.ucl.ac.uk/spm).

#### Regions and measures of interest

Desikan atlas-based ROIs were defined on native-space MRIs using Freesurfer 5.3 and applied to all modalities. The isthmus cingulate cortex was used as the RSC ROI and the inferior parietal cortex as IP. The composite MTL region included the hippocampus, amygdala, entorhinal cortex, and parahippocampal cortex. MRI measures of interest include cortical thickness for cortical ROIs and volume divided by total intracranial volume as estimated by FreeSurfer for the MTL ROI. Analyses using amyloid-PET regional values within the ADNI cohort are performed separately for each tracer because the Centiloid transformation has been validated for global, but not regional, amyloid-PET values from different tracers. Our primary analyses are performed within the FBP subsample because it is larger (n=87); results within the FBB subsample (n=60) are reported in Supplementary Fig. 5. For template-space PET analyses, native-space SUVR images were warped to Montreal Neurological Institute (MNI) template-space following deformation parameters defined on respective structural MRIs using SPM12. Centiloids were calculated using equations validated according to established protocols.^56^ Importantly, FDG-PET and MRI measures were reversed so that greater values indicated greater pathology or neurodegeneration for all imaging modalities.

To define regions connected to RSC and IP, we first determined MNI coordinates for RSC and IP, defined as the voxel of greatest auto-correlation with native-space RSC and IP FDG-PET values within an FDG-PET control group using voxelwise regression analyses (Supplementary Methods). We then used these coordinates on *neurosynth*.*org* to obtain maps of functional connectivity with RSC or IP. The resulting maps were downloaded from Neurosynth and masked with an in-house GM mask then binarized at the 90^th^ percentile of connectivity to create ROIs of most connected regions, referred to as funcROIs moving forward. The resulting funcROIs are groups of statistically defined voxels and do not necessarily correspond to specific neuroanatomical labels. Finally, the discrete cluster corresponding to auto-correlation with RSC or IP was manually removed (see Supplementary Fig. 1 for the funcROI creation process and Figure 1B for an illustration of the final funcROIs). We used mean SUVR values within the funcROI from template-space amyloid- and FTP-PET images as measures of pathology in connected regions.

#### Summary mean images

We created mean summary maps to visualize the patterns of imaging abnormalities in each cohort (Figure 2). FTP- and amyloid-PET abnormalities are shown in mean SUVR units because any elevated cortical signal can be considered pathological. FDG-PET and MRI summary images are shown as statistical maps (deviation from normal controls) because they are more easily interpreted. For these statistical maps, we used W-scores (W, covariate-adjusted z-scores), as described elsewhere.^5,57–59^ FDG-PET W-scores were adjusted for age, and MRI W-scores were adjusted for both age and total intracranial volume (Supplementary Methods). The similarity between imaging abnormality patterns observed in UCSF and ADNI was quantified using a voxelwise spatial correlation approach^21,60,61^; correlations were visualized using a hex scatter plot in MATLAB Version 2015a^62^ and performed within a cortical gray matter mask. For these analyses, r values are interpreted qualitatively: given that correlations were based on 252,753 cortical voxels, p values are irrelevant.

**Figure 2.**
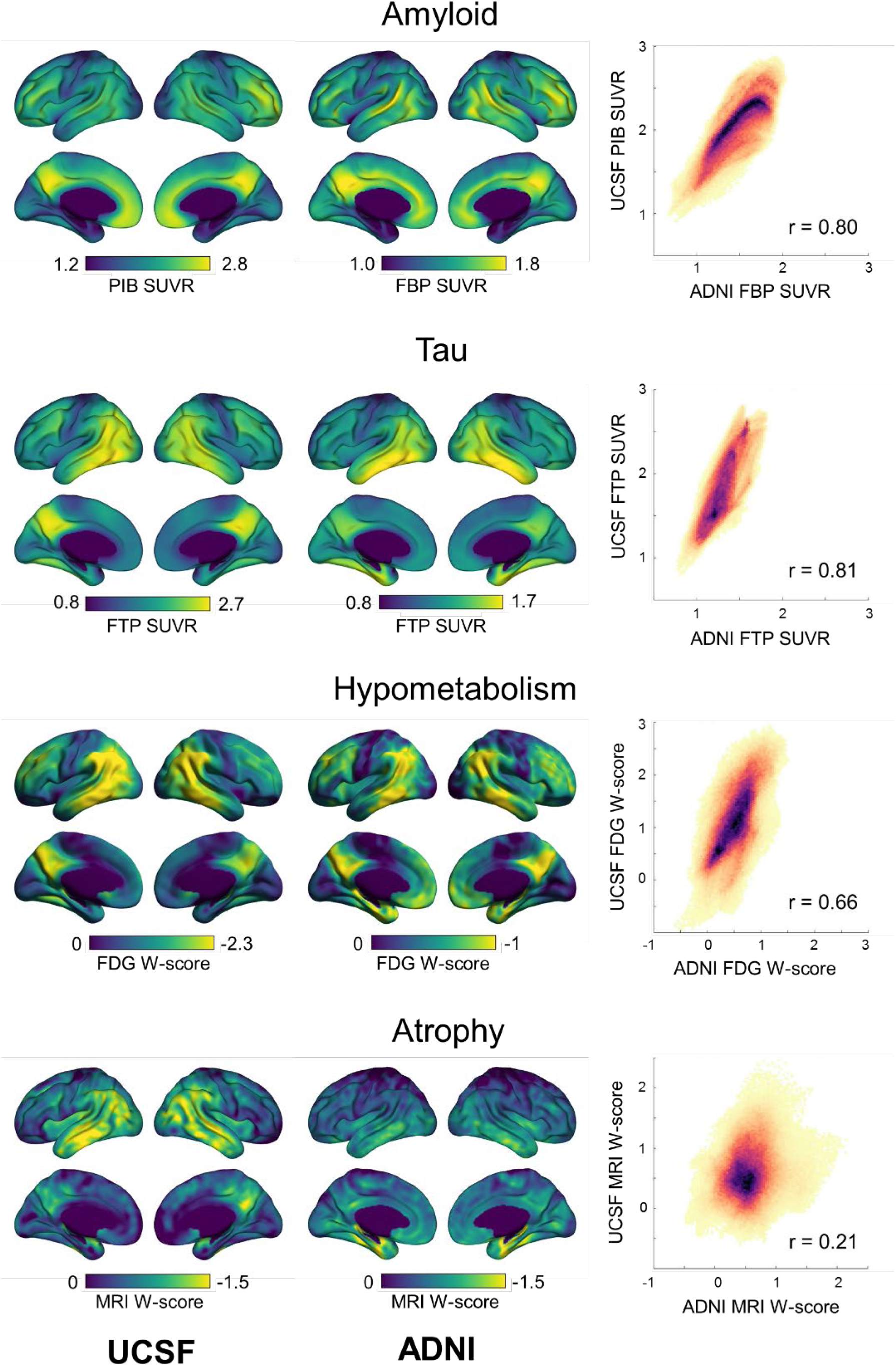
Summaries of imaging modalities, presented as voxelwise means within each cohort. Amyloid- and FTP-PET are presented in SUVR units and FDG-PET and MRI volume in age-adjusted z-score, or W-score, units compared to cognitively normal controls. The scales were adapted for each cohort and modality. Scatter plots (right panel) display a voxel-by-voxel correlation between cohort mean images, with ADNI mean modality values represented on the x-axis and UCSF on the y-axis. All voxels from a cortical gray matter mask are included. Color represents the density of represented voxels. Higher r values indicate greater spatial similarity between mean cohort images.

#### Partial volume correction

To complement our main analyses, we applied a 2-compartment partial volume correction to FDG-PET and repeated the analyses.^63^ The applied gray and white matter mask included voxels with a gray matter or white matter probability of >0.5 according to SPM12 and excluded voxels that were parcellated as a ventricle by FreeSurfer 5.3.

### Statistical analysis

Demographics across cohorts were compared using a standard t-test or Chi-squared test of association where appropriate. To assess the individual relationships between variables of interest and FDG SUVR in RSC and IP, we used bivariate correlations and partial correlations controlling for disease severity and age. We then used linear regression models to assess the independent contributions of these factors with FDG SUVR in the RSC or IP as the dependent variable, based on prior hypotheses. Regression model fit was evaluated using Bayesian information criteria (BIC). Our disease severity measure is a combination of the CDR sum of boxes (CDR-SB) and MMSE scores. Briefly, each measure is z-scored within cohort then averaged within patient to obtain the disease severity score (Supplementary Fig. 2). We used 2-tailed tests for all analyses and an alpha level of .05 to determine significance. Statistical analyses were performed using Jamovi (version 1.1, www.jamovi.org) and R (version 4.0.2, www.R-project.org).

### Data availability statement

The data that support the findings of this study are available from the corresponding author upon request (memory.ucsf.edu/researchtrials/professional/open-science). Gray matter-masked mean images used in Figure 2 are available on Neurovault (neurovault.org/collections/QNGOIQGC/).

## Results

### UCSF participants were significantly younger and more impaired than ADNI participants

We included two independent cohorts of amyloid-positive participants with Mild Cognitive Impairment or dementia: 85 participants from UCSF and 147 from the ADNI study. A summary of demographic characteristics and comparisons between groups is available in Table 1. Briefly, the UCSF cohort was 10.1 years younger (*t*(230) = 8.8, *p* < .001, *d* = 1.20) and more clinically impaired on CDR (*t*(230) = −5.2, *p* < .001, *d* = −0.70 for CDR-SB) and MMSE (*t*(230) = 7.4, *p* < .001, *d* = 1.01) scores. To account for these differences, we included age and disease severity as covariates in our analyses. Years of education and the prevalence of the *APOE* ε4 allele significantly differed, where ADNI had a higher proportion of *APOE* ε4 carriers (68% versus 54%, *χ*^*2*^(1) = 4.2, *p* = .04) and 0.8 fewer years of education (*p* = .02, *d* = −0.31). Sex did not differ between cohorts (*χ*^*2*^(1) = 0.5, *p* = .46). Global amyloid burden as measured by the Centiloid scale^56^ differed such that the UCSF amyloid burden was 12 Centiloids higher (*t*(230) = −2.5, *p* = .01, *d* = −0.34) compared to ADNI.

**Table 1.** Demographic summary and cohort comparison. Continuous variables are shown as mean (standard deviation). For comparisons between cohorts, χ^2^ tests of association were used for discrete variables (Cramer’s V as effect size), Mann-Whitney tests were used for ordinal variables (Cohen’s d as effect size) and t-tests were used for continuous variables (Cohen’s d as effect size). CDR = Clinical Dementia Rating, CDR-SB = Sum of Boxes score, MMSE = Mini Mental State Exam, FBP = ^18^F-Florbetapir, FBB = ^18^F=Florbetaben, PIB = ^11^C-Pittsburgh compound-B.

|  | UCSF<br>(n=85) | ADNI<br>(n=147) | Comparison |  |
| --- | --- | --- | --- | --- |
|  |  |  | Effect size | p |
| **Age** | 65.2 (9.6) | 75.3 (7.8) | d = 1.2 | <.001 |
| **Sex (% F)** | 51% | 46% | V = 0.05 | .46 |
| **Education** | 16.7 (2.3) | 15.9 (2.6) | d = -0.31 | .02 |
| **CDR-Global (% ≥1)** | 47% | 21% | V = 0.27 | <.001 |
| **CDR-SB** | 4.26 (2.2) | 2.74 (2.1) | d = -0.70 | <.001 |
| **MMSE** | 21.2 (6.2) | 25.8 (3.4) | d = 1.01 | <.001 |
| **APOE ε4 carrier (% , missing N)** | 53%, 9 | 68%, 23 | V = 0.15 | .03 |
| **Amyloid-PET burden (Centiloids)** | 98 (30) | 86 (39) | d = -0.34 | .01 |
| **Amyloid tracer (FBP/FBB/PIB)** | 0/0/85 | 87/60/0 | – | – |

Demographic comparisons between amyloid-PET tracer subgroups in the ADNI cohort are presented in Supplementary Table 3. The FBB subgroup was slightly younger (74.5 years old versus 76.3 years old in FBP subgroup, *p* = .06) and had a higher proportion of *APOE* ε4 carriers (80% versus 60% in FBP subgroup, *p* = .02). Statistical models for ADNI that include amyloid-PET measures are performed within the FBP subgroup; all other analyses are performed in the whole group.

### Regions of imaging abnormalities were comparable between cohorts though intensity differed

A visual summary of imaging abnormalities between cohorts is available in Figure 2. Qualitatively, FDG- and FTP-PET abnormalities followed a canonical temporo-parietal-predominant pattern with involvement of dorsolateral prefrontal regions in both cohorts. Cohorts differed primarily on the magnitude of abnormalities, with greater hypometabolism and tau pathology in UCSF. The pattern of gray matter atrophy was temporo-parietal-predominant in the UCSF cohort and MTL-predominant in the ADNI cohort. On amyloid-PET, a diffuse neocortical pattern was noted in both cohorts.

Similarities in imaging patterns across cohorts were also quantified using voxelwise spatial correlations (Figure 2, right panel). Patterns of amyloid-PET and tau-PET were highly similar across cohorts (*r* = .80 and .81, respectively). Regarding neurodegeneration biomarkers, patterns of hypometabolism observed in ADNI and UCSF were strongly correlated (*r* = .66) while patterns of gray matter atrophy showed a weaker spatial similarity (*r* = .21).

Spatial correlations were also investigated across imaging modalities in each cohort separately (Supplementary Fig. 3). Briefly, average patterns of hypometabolism were more similar to tau-PET patterns (*r* = .76 in UCSF, *r* = .55 in ADNI) than to amyloid-PET (*r* = .28 in UCSF, *r* = .22 in ADNI) or atrophy (*r* = .58 in UCSF, *r* = .38 in ADNI) patterns. In both cohorts, the spatial correlation between hypometabolism and tau patterns was the strongest of all pairwise associations and exceeded the similarity between tau and atrophy (*r* = .44 in UCSF, *r* = .38 in ADNI).

**Figure 3.**
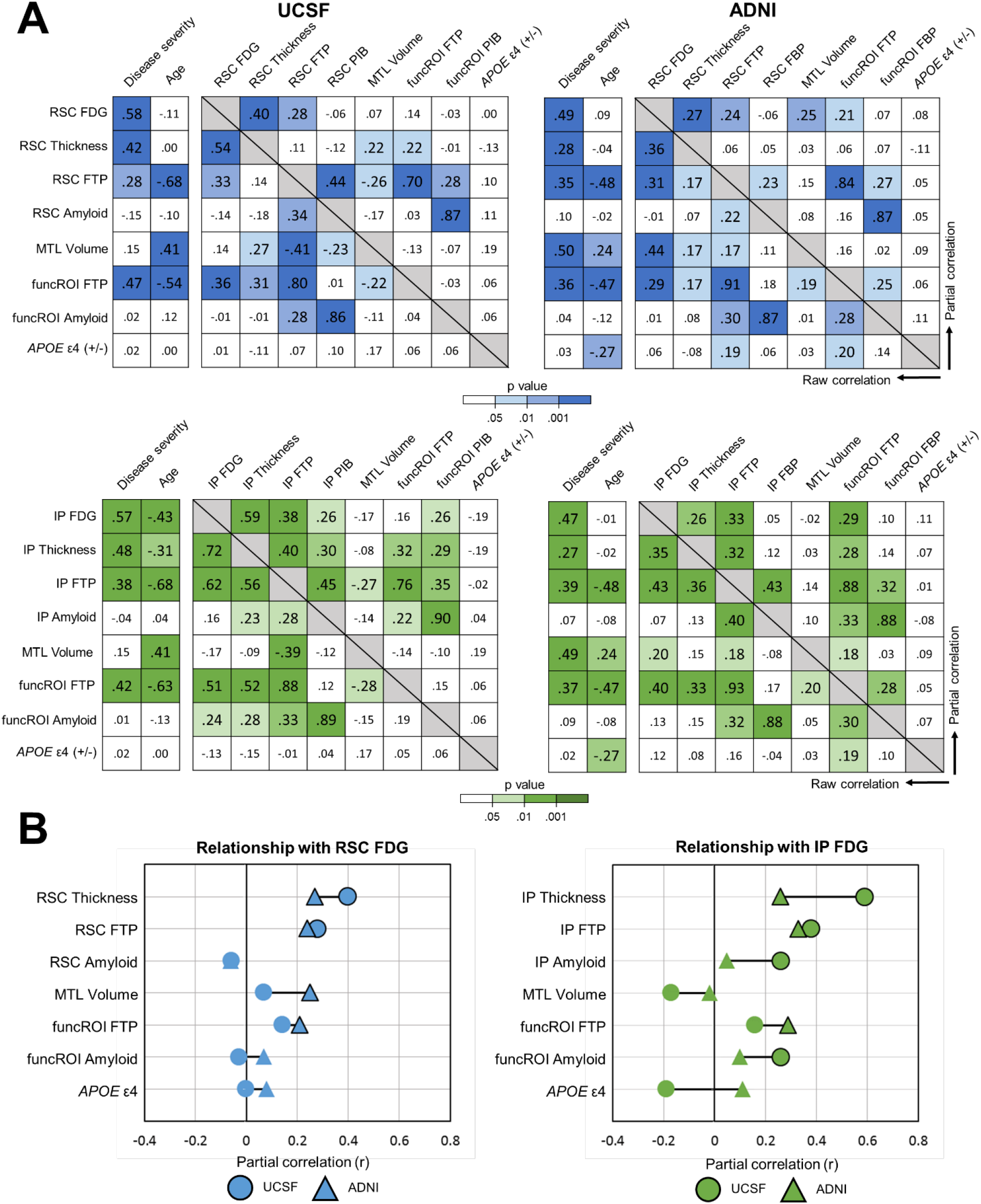
Correlations across measures of interest. (**A**) Correlation matrices with bivariate correlations in the bottom left portion and partial correlations controlling for age and disease severity in the top right. Color saturation corresponds to p-value. (**B**) An alternative presentation of partial correlations with FDG in RSC or IP with a direct visual comparison between ADNI (triangles) and UCSF (circle) cohorts. The presence of shape border reflects statistical significance defined as p<.05. MTL volume was divided by total intracranial volume before analyses.

### FTP SUVR and local thickness consistently correlate with RSC and IP FDG

To illustrate individual associations between our hypothesized measures and FDG SUVR in RSC and IP, we used correlation analyses, summarized in Figure 3. Bivariate correlations differed by region and cohort, with FTP SUVR and local thickness displaying the most consistent relationship with regional FDG SUVR across regions and cohorts (*r*s > .31, *p*s < .01). *APOE* ε4 was consistently unrelated to regional FDG SUVR (absolute *r*s < .15, *p*s > .05).

Using partial correlations controlling for age and disease severity, associations with local FTP SUVR and thickness remained significant across regions and cohorts (*r*s = .24 to .38 for FTP, *r*s = .26 to .59 for thickness; all *p*s < .01). FDG SUVR was often related to FTP SUVR in connected regions (i.e. funcROI FTP), though to a lower magnitude than local measures (*r*s = .14 to .29, *p*s = <.001 to .22). Partial correlations between FDG SUVR and FTP SUVR measures were compared using a one-tailed r-to-z transform to statistically assess whether local correlations were stronger than distant correlations given the collinearity between FTP measures. Local FTP-FDG correlations were significantly stronger than funcROI FTP-FDG correlations in the UCSF (*p* = .046 for RSC, *p* = .001 for IP) but not ADNI (*p* = .25 for RSC, *p* = .15 for IP) cohort.

Of note, sample sizes differ for analyses with amyloid-PET in ADNI (*n* = 87 with FBP) and with *APOE* ε4 (*n* = 82 in UCSF, *n* = 124 in ADNI). Amyloid-PET analyses repeated within the FBB subset of ADNI yielded similar results (Supplementary Fig. 5).

### Primarily local factors are associated with FDG SUVR in linear regression analyses

Next, linear regression models were used to test the relative associations of each predictor with FDG SUVR in our cohorts (see Tables 2-3). Multiple regressions were performed separately for each cohort and region where FDG SUVR in RSC or IP is the singular dependent variable. Each hypothesized factor was added individually to assess if it improved the model overall. Local FTP SUVR and thickness were included in all hypothesis-testing models due to their consistently significant bivariate and partial correlations with FDG SUVR shown previously.

**Table 2.** Linear regression models testing all hypothesized factors, UCSF. Analyses are run within the UCSF cohort and within region, where FDG in either RSC (A) or IP (B) is the singular dependent variable. Local cortical thickness and FTP are included in all hypothesis-testing models due to their robust associations with FDG in previous correlation analyses. Models including *APOE* **ε** 4 are run within a smaller sample (n=76), and the reference levels for R^2^ and BIC are modified accordingly. MTL volume was divided by total intracranial volume prior to analyses. β = standardized estimate; BIC = Bayesian Information Criteria.

| **A. RSC FDG** | **Model 0** |  | **Model 1** |  | **Model 2** |  | **Model 3** |  | **Model 4** |  |
| --- | --- | --- | --- | --- | --- | --- | --- | --- | --- | --- |
| **Predictors** | $\beta$ | p | $\beta$ | p | $\beta$ | p | $\beta$ | p | $\beta$ | p |
| **Disease severity** | 0.60 | <.001 | 0.38 | <.001 | 0.36 | <.001 | 0.44 | <.001 | 0.37 | <.001 |
| **Age** | 0.01 | .88 | 0.15 | .15 | 0.14 | .24 | 0.17 | .13 | 0.20 | .10 |
| **RSC thickness** | - | - | 0.34 | <.001 | 0.32 | <.001 | 0.37 | <.001 | 0.32 | .001 |
| **RSC FTP** | - | - | 0.28 | .02 | 0.30 | .01 | 0.59 | .001 | 0.34 | .007 |
| **MTL volume** | - | - | - | - | 0.06 | .52 | - | - | - | - |
| **funcROI FTP** | - | - | - | - | - | - | -0.34 | .03 | - | - |
| **funcROI PIB** | - | - | - | - | - | - | -0.15 | .10 | - | - |
| **APOE ε4** | - | - | - | - | - | - | - | - | 0.02 | .90 |
| **R<sup>2</sup> (ΔR<sup>2</sup>)** | 0.33 (Ref) |  | 0.47 (0.15) |  | 0.47 (0.15) |  | 0.51 (0.18) |  | 0.51 (0.15) |  |
| **BIC (ΔBIC<sub>0</sub>)** | -103 (Ref) |  | -114 (-11) |  | -110 (-7) |  | -111 (-8) |  | -110 (-7) |  |

**Table 2. Linear regression models testing all hypothesized factors, UCSF.**
| **B. IP FDG** | **Model 0** |  | **Model 1** |  | **Model 2** |  | **Model 3** |  | **Model 4** |  |
| --- | --- | --- | --- | --- | --- | --- | --- | --- | --- | --- |
| **Predictors** | $\beta$ | p | $\beta$ | p | $\beta$ | p | $\beta$ | p | $\beta$ | p |
| **Disease severity** | 0.52 | <.001 | 0.26 | <.001 | 0.28 | <.001 | 0.30 | <.001 | 0.23 | .009 |
| **Age** | -0.34 | <.001 | -0.12 | .20 | -0.10 | .28 | -0.15 | .11 | -0.12 | .23 |
| **IP thickness** | - | - | 0.46 | <.001 | 0.46 | <.001 | 0.46 | <.001 | 0.47 | <.001 |
| **IP FTP** | - | - | 0.19 | .08 | 0.16 | .14 | 0.45 | .006 | 0.17 | .16 |
| **MTL volume** | - | - | - | - | -0.07 | .37 | - | - | - | - |
| **funcROI FTP** | - | - | - | - | - | - | -0.34 | .02 | - | - |
| **funcROI PIB** | - | - | - | - | - | - | 0.01 | .88 | - | - |
| **APOE ε4** | - | - | - | - | - | - | - | - | -0.12 | .42 |
| **R<sup>2</sup> (ΔR<sup>2</sup>)** | 0.44 (Ref) |  | 0.65 (0.21) |  | 0.65 (0.21) |  | 0.67 (0.23) |  | 0.65 (0.22) |  |
| **BIC (ΔBIC<sub>0</sub>)** | -96 (Ref) |  | -126 (-30) |  | -123 (-27) |  | -124 (-28) |  | -120 (-24) |  |

**Table 3.** Linear regression models testing all hypothesized factors, ADNI. Analyses are run within the ADNI cohort and within region, where FDG in either RSC (A) or IP (B) is the singular dependent variable. Local cortical thickness and FTP are included in all hypothesis-testing models due to their robust associations with FDG in previous correlation analyses. Models including funcROI FBP or *APOE* **ε** 4 are run within smaller samples (n=87 for Model 3 and n=124 for Model 4), and the reference levels for R^2^ and BIC are modified accordingly. MTL volume was divided by total intracranial volume prior to analyses. β = standardized estimate; BIC = Bayesian Information Criteria.

| **A. RSC FDG** | **Model 0** |  | **Model 1** |  | **Model 2** |  | **Model 3** |  | **Model 4** |  |
| --- | --- | --- | --- | --- | --- | --- | --- | --- | --- | --- |
| **Predictors** | $\beta$ | p | $\beta$ | p | $\beta$ | p | $\beta$ | p | $\beta$ | p |
| **Disease severity** | 0.48 | <.001 | 0.32 | <.001 | 0.23 | .007 | 0.26 | .02 | 0.29 | <.001 |
| **Age** | 0.05 | .47 | 0.19 | .02 | 0.13 | .11 | 0.27 | .03 | 0.28 | .003 |
| **RSC thickness** | - | - | 0.24 | .001 | 0.23 | .001 | 0.20 | .04 | 0.26 | .001 |
| **RSC FTP** | - | - | 0.25 | .005 | 0.22 | .01 | 0.34 | .13 | 0.27 | .005 |
| **MTL volume** | - | - | - | - | 0.22 | .008 | - | - | - | - |
| **funcROI FTP** | - | - | - | - | - | - | -0.09 | .65 | - | - |
| **funcROI FBP** | - | - | - | - | - | - | 0.00 | .96 | - | - |
| **APOE ε4** | - | - | - | - | - | - | - | - | 0.20 | .23 |
| **R<sup>2</sup> (ΔR<sup>2</sup>)** | 0.24 (Ref) |  | 0.33 (0.09) |  | 0.37 (0.13) |  | 0.28 (0.09) |  | 0.36 (0.13) |  |
| **BIC (ΔBIC<sub>0</sub>)** | -121 (Ref) |  | -130 (-9) |  | -133 (-12) |  | -113 (+8) |  | -129 (-8) |  |

**Table 3. Linear regression models testing all hypothesized factors, ADNI.**
| **B. IP FDG** | **Model 0** |  | **Model 1** |  | **Model 2** |  | **Model 3** |  | **Model 4** |  |
| --- | --- | --- | --- | --- | --- | --- | --- | --- | --- | --- |
| **Predictors** | $\beta$ | p | $\beta$ | p | $\beta$ | p | $\beta$ | p | $\beta$ | p |
| **Disease severity** | 0.47 | <.001 | 0.29 | <.001 | 0.32 | <.001 | 0.28 | .02 | 0.28 | .001 |
| **Age** | -0.05 | .48 | 0.12 | .17 | 0.14 | .12 | 0.16 | .20 | 0.16 | .08 |
| **IP thickness** | - | - | 0.16 | .04 | 0.16 | .04 | 0.10 | .34 | 0.15 | .07 |
| **IP FTP** | - | - | 0.32 | .001 | 0.33 | <.001 | 0.34 | .26 | 0.36 | <.001 |
| **MTL volume** | - | - | - | - | -0.07 | .38 | - | - | - | - |
| **funcROI FTP** | - | - | - | - | - | - | -0.05 | .86 | - | - |
| **funcROI FBP** | - | - | - | - | - | - | 0.01 | .90 | - | - |
| **APOE ε4** | - | - | - | - | - | - | - | - | 0.19 | .24 |
| **R<sup>2</sup> (ΔR<sup>2</sup>)** | 0.22 (Ref) |  | 0.33 (0.11) |  | 0.33 (0.11) |  | 0.26 (0.07) |  | 0.37 (0.13) |  |
| **BIC (ΔBIC<sub>0</sub>)** | -134 (Ref) |  | -146 (-12) |  | -141 (-7) |  | -125 (+9) |  | -143 (-9) |  |

The addition of local FTP SUVR and thickness measures (local model) significantly improved models containing only age and disease severity (Δ*R*^*2*^ = .09 to .21, Δ*BIC* = −9 to −30), with each local factor contributing significantly and independently (*p*s = .08 to <.001, *β* = 0.16 to 0.46). The addition of MTL volume significantly improved this local model only when predicting RSC FDG-SUVR within ADNI (*p* = .008, Δ*R*^*2*^ = .04 and Δ*BIC* = −3 compared to local). When using BIC to identify the best model, the local model performed optimally for both RSC and IP in UCSF and IP in ADNI. A model including local factors and pathology in the funcROIs performed well but not optimally within the UCSF cohort, though associations with pathology in connected regions appeared in a negative direction. This flip in sign may have occurred due to collinearity between FTP measures in local and functionally connected regions, as has been described previously.^64^

### Complementary analyses on the association between MTL volume and RSC metabolism

The main discrepancy in the analyses presented above regarded the independent effect of MTL volume on RSC FDG SUVR, which was significant in ADNI (*p* = .008), but not UCSF (*p* = .52). We performed exploratory interaction analyses within cohort assessing whether age or disease severity (i.e. the two main factors that differ between the cohorts) were modulating the relationship between MTL volume and RSC FDG SUVR. We did not find an interaction between MTL volume and age on RSC FDG SUVR (*p* = .50 in UCSF, *p* = .58 in ADNI). In contrast, a disease severity * MTL volume interaction was found in both cohorts such that the relationship between MTL volume and RSC FDG SUVR was more positive at earlier disease stages (Figure 4; *t* = −1.9, *p* = .06 in UCSF; *t* = −2.1, *p* = .046 in ADNI).

**Figure 4.**
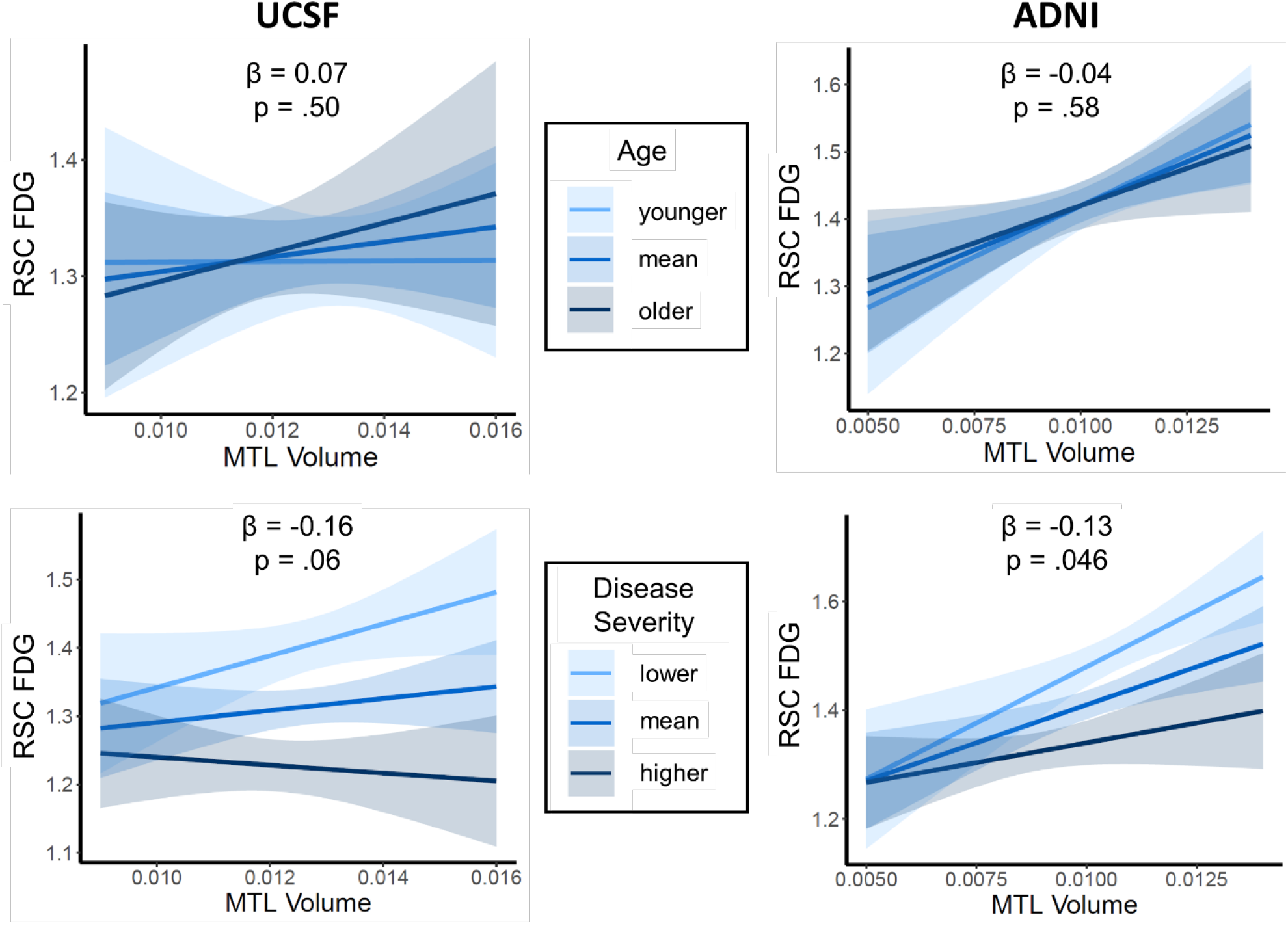
Interaction analyses between MTL volume and age or disease severity in predicting RSC hypometabolism. Each plot represents a separate model. Models include age, disease severity, MTL volume (divided by total intracranial volume), and the interaction term between MTL volume and age or disease severity. Reported p and β (standardized estimate) values refer to the interaction term. Bins refer to mean ± 1 standard deviation of within-cohort age or disease severity. For age, these values correspond to 55, 65, and 74 years for UCSF; or 67, 75, and 83 years for ADNI. For disease severity, bins correspond to CDR-SB/MMSE values of 2.0/27, 4.0/20; and 6.0/14 for UCSF; or 0.5/29, 1.5/24, and 3.5/20 for ADNI.

To assess whether the absence of association between MTL volume and RSC FDG SUVR in the UCSF cohort could be driven by the inclusion of non-amnestic phenotypes, we repeated the analyses in the UCSF cohort after excluding patients with the logopenic variant of primary progressive aphasia or posterior cortical atrophy. Results in this subset mirrored those in the whole group, where MTL volume and RSC FDG SUVR were not significantly related in both bivariate correlations (*r* = .06, *p* = .63) and linear regression analyses (equivalent to Model 2 in Table 2A, *p* = .91 for MTL).

### Results using partial volume-corrected data

Using FDG partial volume-corrected data, many results remained but were somewhat attenuated. In the partial correlations controlling for age and disease severity, relationships with local thickness remained in the UCSF cohort (*p* = .01 for RSC, *p* = .001 for IP), but not the ADNI cohort (*p* = .20 for RSC, *p* = .58 for IP). Relationships with local FTP were attenuated (*p*s = .04 – .36). The relationship between RSC FDG SUVR and MTL volume in the ADNI cohort remained (*r* = .18, *p* = .03; Supplementary Fig. 4).

## Discussion

In two independent cohorts of symptomatic patients on the Alzheimer’s continuum, we found that decreased glucose metabolism as measured by FDG-PET associates with local atrophy and tau pathology, with medial temporal atrophy associating at early clinical stages. Molecular pathology in functionally connected regions and the presence of the *APOE* ε4 allele did not significantly contribute to hypometabolism after accounting for local factors.

Decreased FDG-PET consistently correlated locally with decreased cortical thickness in the RSC and IP, which is unsurprising given that both are measures of neurodegeneration; this relationship has been reported many times previously.^5,17,18,65^ However, FDG-PET and structural MRI-derived measures are not redundant,^5,15,16,66,67^ as is illustrated by the discordant patterns of modality abnormalities in the present cohorts (Figure 2): while patterns of decreased FDG-PET followed a similar temporo-parietal pattern across the two cohorts, decreased gray matter volume was most prominent in the medial (ADNI) or lateral (UCSF) temporal lobes. Given the inconsistencies between cohorts in MRI patterns in contrast with FDG-PET patterns, FDG-PET may be a more reliable measure for detecting an Alzheimer’s disease-associated pattern of neurodegeneration. The RSC and IP, while among the most hypometabolic regions, were not always the most atrophic. This discrepancy cannot be explained by one modality simply being more sensitive than the other, though sensitivity to neurodegeneration may differ by region as well as modality.^17,18,68^ In some neurodegenerative diseases, MRI and FDG-PET abnormalities are highly consistent; the discrepancy between these modalities appears to be the most salient in Alzheimer’s disease.^21^ These findings suggest that measures of glucose metabolism in Alzheimer’s disease capture processes beyond local atrophy, as has been postulated previously.^18,21^

Tau-PET correlated locally with decreased FDG-PET independent of co-occurring cortical thinning. This finding agrees with existing literature that links tau aggregation to metabolic dysfunction, as evidenced by the consistent co-localization and correlation between tau-PET and decreased FDG-PET, especially in groups of amyloid-positive individuals like the cohorts presented here.^4,18,22,24,26,27^ Given that FDG-PET primarily captures synaptic activity ^2^, tau pathology may disrupt synaptic function^69,70^ and thus reduce regional glucose uptake independently of gross structural change. Additionally, previous work in our lab found that tau-PET topography and magnitude are more predictive of longitudinal change in cortical thickness than cross-sectional cortical thickness.^60^ It has been postulated that FDG-PET as a biomarker becomes abnormal earlier than MRI in Alzheimer’s disease^17,71–74^ and other neurodegenerative disorders.^68,75^ Taken together, these findings suggest that tau pathology may be more closely linked temporally to metabolic than structural abnormalities. If so, the stronger cross-sectional relationship observed between tau and hypometabolism than between tau and atrophy (Figure 3 and Supplementary Fig. 3) may reflect a tighter temporal link.

We did not find an effect of local amyloid-PET on FDG-PET, which is consistent with other cohorts of amyloid-positive individuals.^5,31–35^ Given that the degree of amyloid pathology reaches a plateau soon after symptom onset^76^ in contrast with the increasing levels of tau pathology,^77,78^ it is likely that any direct influence from amyloid plaque pathology may be difficult to detect given the neurotoxicity of tau pathology at this symptomatic stage.

Lower MTL volume was related to decreased FDG-PET signal in the RSC, but not IP. This regional specificity has been reported previously with hypotheses regarding the underlying mechanisms.^28,39^ For one, the RSC, in contrast to the IP, is proximal to MTL regions, especially the parahippocampal cortex, so this relationship could reflect neurodegeneration of neighboring structures.^79^ The spatial extent of FDG-PET abnormalities is larger than that of structural MRI,^22^ so the observed effect could represent a hypometabolism “halo” surrounding adjacent regions of atrophy. Alternatively, MTL atrophy could result in degeneration of the cingulum bundle, deafferentation of the RSC, and thus decreased postsynaptic activity as measured by FDG-PET.^18,28,38,39,65,80^ The MTL is structurally well-connected to the RSC, but not IP,^51^ and hypometabolism has been linked to impaired white matter tract integrity in cognitively impaired patients,^35,39,80,81^ providing evidence for this mechanism.

The relationship between MTL atrophy and RSC FDG-PET was significant only in the ADNI cohort. This cohort specificity could be due to the intrinsic differences in the characteristics of the samples, notably differences in age, disease severity, or inclusion criteria: the ADNI patients were older, less severely impaired, and included patients with more amnestic-predominant presentations than the UCSF participants. Converging evidence suggests that in older patients with pathology-proven Alzheimer’s disease, MTL atrophy is related to both Alzheimer’s disease tau pathology and frequently comorbid TDP-43 pathology,^82–84^ an entity also known as limbic-predominant age-related TDP-43 encephalopathy^85^ and often associated with hippocampal sclerosis. In the older ADNI cohort, MTL degeneration could therefore stem not only from Alzheimer’s disease neuropathology but also from comorbid TDP-43 or hippocampal sclerosis.^86^ However, we did not find evidence that age modulates the relationship between lower MTL volume and RSC hypometabolism (Figure 4), arguing against the hypothesis that older age was the primary driver of the association seen in ADNI. Each cohort was limited in its age range, though, and a broader cohort may be better suited to addressing this question. In addition, the exclusion of non-amnestic clinical phenotypes included in the UCSF cohort did not affect our results and could not explain the distinct patterns observed across cohorts.

In contrast, interaction analyses (Figure 4) showed that the relationship between MTL atrophy and RSC hypometabolism was modulated by disease severity, such that it was more positive at milder levels of impairment. Alzheimer’s disease is sometimes referred to as a “disconnection syndrome”, where functional and structural connectivity across neural networks are increasingly disrupted over the course of the disease.^87,88^ The relationship between MTL atrophy and RSC hypometabolism could be particularly significant at earlier disease stages because the MTL and RSC become increasingly disconnected over time, affecting disease mechanisms.^89^ This disconnection hypothesis may also explain the lack of an association between regional hypometabolism and molecular pathology in distant but functionally connected regions. Many studies observing this relationship were performed in preclinical populations when network connectivity is relatively robust and may better facilitate disease processes.^40^ Also at early disease stages, molecular pathology burden, especially tau, is spatially restricted,^90^ whereas at later stages it becomes more widespread and more spatially homogeneous, with stronger inter-region correlations within amyloid or tau measures.^24^ Such collinearity affects the statistical power required to detect region-specific relationships. Nonetheless, these findings conflict with the study by Pascoal *et al*.,^34^ who recently found that amyloid pathology as measured by amyloid-PET in distant regions, but not locally, may induce regional hypometabolism via functional connections, even in patients with Mild Cognitive Impairment. However, this study included phosphorylated tau in the cerebrospinal fluid (CSF p-tau) as a measure of tau pathology, while the present study uses tau-PET. CSF p-tau only moderately correlates with tau-PET, and tau-PET is more closely related to cognitive decline and neurodegeneration than CSF p-tau, so these measures likely capture different processes.^91,92^ It is therefore possible that the reported association between remote amyloid and hypometabolism could still be mediated by tau pathology in humans.

We did not see a relationship between hypometabolism and the presence of the *APOE* ε4 allele. This finding is largely consistent with existing literature, where this relationship is more often found in asymptomatic rather than symptomatic individuals.^30,31,46^ Previous studies that have observed this relationship in clinical cohorts did not include tau-PET and either included amyloid-negative individuals or a distinction between *APOE* ε4 heterozygotes and homozygotes.^47,48^ Additionally, the *APOE* ε4-associated decrease in glucose metabolism in Alzheimer’s disease-associated regions observed in clinically normal cohorts is small compared to metabolic decreases due to clinical neurodegeneration.^46^ Therefore, this small effect may be masked by clinically relevant processes at this stage of the disease.

A strength of the present study is the inclusion of two relatively large complementary samples of biomarker-supported Alzheimer’s disease patients. With complementary cohorts, we were able to test our hypothesized factors across a wider range of ages and levels of impairment, allowing for greater generalizability to Alzheimer’s disease populations. Our results were consistent across these clinical differences as well as differences in study inclusion criteria, site, and scanner. Additionally, we assessed multiple factors simultaneously to directly compare the relative contributions of each factor and performed analyses both with and without partial volume correction. However, the present study is cross-sectional in design, so findings of distinct relationships at different disease stages should be confirmed in a longitudinal design. Our findings also may not apply to preclinical stages, as our cohorts consisted of only symptomatic Alzheimer’s disease patients, or to the more diverse populations that are represented in memory clinics, given the high educational attainment and the high proportion of non-Hispanic White individuals included in the cohorts. Finally, while we assessed multiple factors, there are many other possible determinants of hypometabolism that were not addressed in the current study, including but not limited to white matter degeneration,^35^ local inflammation,^93^ and vascular changes.^81^

In conclusion, we found that posterior cortical hypometabolism in Alzheimer’s disease reflects primarily local atrophy and tau pathology of the possible factors tested. Medial temporal atrophy is related to RSC hypometabolism only at early disease stages. Our data also showed that molecular pathology in remote brain regions and the presence of the *APOE* ε4 allele may not be related to a greater degree of hypometabolism at the symptomatic stage of Alzheimer’s disease. These results suggest that Alzheimer’s disease hypometabolism is primarily influenced by neurodegeneration and tau, but not amyloid, pathology, reflecting a tau-centric mechanism of synaptic dysfunction. Given the added value of non-atrophy measures in predicting hypometabolism, FDG-PET and MRI may not be interchangeable measures of Alzheimer’s-related neurodegeneration.

## Acknowledgments

We thank patients and families for their commitment.

Avid Radiopharmaceuticals enabled the use of the [^18^F]Flortaucipir tracer by providing precursor, but did not provide direct funding and was not involved in data analysis or interpretation.

## Funding

The present study was supported by the National Institutes of Health/National Institute of Aging grants NIH/NIA P50-AG23501 (to GDR, BLM), UCSF ADRC P50-AG023501, P30-AG062422 (to BLM, GDR), P01-AG019724 (to BLM), R01-AG045611 (to GDR), R01-AG034570 (to WJJ), R01-AG032306 (to HLR), NIH/NINDS R01-NS050915 (to MLG), K99AG065501 (to RLJ), K24-AG053435 (to HJR), Rainwater Charitable Foundation (to GDR, WJJ), Alzheimer’s Association (to RLJ, AARF:16-443577 and DSM, AACSF:19-617663).

Data collection and sharing for this project were funded by the Alzheimer’s Disease Neuroimaging Initiative (ADNI) (National Institutes of Health Grant U01 AG024904) and DOD ADNI (Department of Defense award number W81XWH-12-2-0012). ADNI is funded by the National Institute on Aging, the National Institute of Biomedical Imaging and Bioengineering, and through generous contributions from the following: AbbVie, Alzheimer’s Association; Alzheimer’s Drug Discovery Foundation; Araclon Biotech; BioClinica, Inc.; Biogen; Bristol-Myers Squibb Company; CereSpir, Inc.; Cogstate; Eisai Inc.; Elan Pharmaceuticals, Inc.; Eli Lilly and Company; EuroImmun; F. Hoffmann-La Roche Ltd and its affiliated company Genentech, Inc.; Fujirebio; GE Healthcare; IXICO Ltd.; Janssen Alzheimer Immunotherapy Research & Development, LLC.; Johnson & Johnson Pharmaceutical Research & Development LLC.; Lumosity; Lundbeck; Merck & Co., Inc.; Meso Scale Diagnostics, LLC.; NeuroRx Research; Neurotrack Technologies; Novartis Pharmaceuticals Corporation; Pfizer Inc.; Piramal Imaging; Servier; Takeda Pharmaceutical Company; and Transition Therapeutics. The Canadian Institutes of Health Research is providing funds to support ADNI clinical sites in Canada. Private sector contributions are facilitated by the Foundation for the National Institutes of Health (www.fnih.org). The grantee organization is the Northern California Institute for Research and Education, and the study is coordinated by the Alzheimer’s Therapeutic Research Institute at the University of Southern California. ADNI data are disseminated by the Laboratory for Neuro Imaging at the University of Southern California.

## Competing interests

Amelia Strom, Leonardo Iaccarino, Lauren Edwards, Orit Lesman-Segev, David Soleimani-Meigooni, Julie Pham, Susan Landau, Howard J. Rosen, Maria Luisa Gorno-Tempini, Renaud La Joie have nothing to disclose. Suzanne L. Baker consults for Genentech. Bruce L Miller receives research support from the NIH/NIA and the Centers for Medicare & Medicaid Services (CMS) as grants for the Memory and Aging Center. As an additional disclosure, Dr. Miller serves as Medical Director for the John Douglas French Foundation; Scientific Director for the Tau Consortium; Director/Medical Advisory Board of the Larry L. Hillblom Foundation; Scientific Advisory Board Member for the National Institute for Health Research Cambridge Biomedical Research Centre and its subunit, the Biomedical Research Unit in Dementia (UK); and Board Member for the American Brain Foundation (ABF). William J Jagust has served as a consultant to BioClinica, Genentech, and Novartis Pharmaceuticals. Gil D Rabinovici receives research support from Avid Radiopharmaceuticals, GE Healthcare, and Life Molecular Imaging, and has received consulting fees or speaking honoraria from Axon Neurosciences, Avid Radiopharmaceuticals, GE Healthcare, Johnson & Johnson, Roche, Eisai, Genentech, Merck. He is an associate editor of JAMA Neurology.

## Abbreviations

*APOE* ε4: apolipoprotein E ε4;
FBP: [^18^F]-Florbetapir;
FDG: [^18^F]-Fluorodeoxyglucose;
FTP: [^18^F]-Flortaucipir;
IP: inferior parietal lobe;
MTL: medial temporal lobe;
PIB: ^11^C-Pittsburgh compound-B;
RSC: retrosplenial cortex;
SUVR: standard uptake value ratio;

