## Supplementary material for "Glucose metabolism reflects local atrophy and tau pathology in symptomatic Alzheimer’s disease"

### Supplementary Methods

#### Creating FDG SUVR images using a pons reference region for ADNI

UCSF patient pons ROIs, which are generated via manual edit of the Freesurfer brainstem parcellation, were warped to MNI space using transformations derived using SPM12 on corresponding MRIs. Corresponding pons files were averaged and binarized at > 0.1 to create a template-space pons reference region. The template pons was then reverse-normalized to each ADNI participant's native space using nearest neighbor interpolation. We multiplied this native-space pons by the participant's white matter probability map binarized at > 0.2 to ensure sufficient adherence to patient anatomy. The output at each step of the process was checked carefully.

Each ADNI participant's mean FDG image was coregistered to their corresponding MRI; mean pons value was extracted from each FDG image using the customized pons file described above to create SUVR images.

#### Creating mean summary images (Figure 1)

To visualize the pattern of PET imaging abnormalities in each cohort, SUVR images were first warped to MNI template space according to SPM-defined deformation parameters obtained based on the respective structural MRI. SPM was then used to calculate within-cohort, within-tracer mean SUVR images for amyloid-PET and FTP-PET. Because group average FDG SUVR images are not easily interpreted visually due to physiological variations in FDG SUVR values across regions, we calculated a z-score controlling for age based on a group of normal controls (NC), referred to as a W-score,[1–4] within each voxel for each patient. Mean W-score images were then created using SPM.

For MRI summary images, we used the Segment tool in SPM12 to create modulated warped gray matter probability maps for each patient and NC. These maps were then smoothed to final PET resolution (8 $mm^3$). W-scores were then calculated within each voxel for each patient based on the NC groups described in Tables S1-S2, controlling for both age and estimated total intracranial volume.

Resulting mean images were rendered on a three-dimensional brain surface using the BrainNet Viewer (www.nitrc.org/projects/bnv/) and default interpolation without adjusting for null. Hex scatter plots were created in Matlab[5] using gray matter-masked mean images for each axis. Perceptually uniform color scales were used (viridis for mean images, inversed magma for hex scatter plots; https://matplotlib.org/).

##### Cognitively normal control groups for UCSF

NC groups were included from the Berkeley Aging Cohort Study (BACS) and Neuroimaging in Frontotemporal Dementia (NIFD) study. No cognitively normal participant had both FDG and MRI on the same scanners as the patients, so NC groups are modality-specific. Control groups did not differ from each other nor the UCSF patient group on age, sex, and education. Details on BACS inclusion criteria can be found in previous publications.[6] For up-to-date information on NIFD participation and protocol, please visit http://memory.ucsf.edu/research/studies/nifd.

| | Patients | NCs | | p |
|---|---|---|---|---|
| | | FDG | MRI | |
| **N** | 85 | 78 | 80 | – |
| **Age** | 64.7 (9.6) | 66.8 (14.7) | 66.6 (17.9) | .59 |
| **Sex (% female)** | 51% | 55% | 56% | .74 |
| **Education** | 16.9 (2.8) | 16.8 (2.1) | 16.7 (2.7) | .59 |
| **MMSE** | 21.2* (6.2) | 28.9 (1.1) | 29.0 (1.1) | <.001 |

**Supplementary Table 1. Group comparison between UCSF patient and NC groups.**

Continuous variables are shown as mean (standard deviation). For comparisons across the three groups, $\chi^2$ tests of association were used for discrete and ordinal variables and ANOVAs were used for continuous variables. *In post-hoc analyses, patient MMSE was lower than each NC group (ps <.001), but the NC groups did not differ from each other (p = .99).

##### Control group for ADNI

A group of age-matched normal controls were selected from the ADNI study based on availability of FDG-PET and MRI data. All had a global CDR of 0 and an MMSE of at least 26. Control groups did not differ from the ADNI patient group on age, sex, and education.

| | Patients | NCs | p |
|---|---|---|---|
| **N** | 147 | 63 | – |
| **Age** | 75.3 (7.8) | 73.2 (6.7) | .06 |
| **Sex (% female)** | 46% | 56% | .18 |
| **Education** | 15.9 (2.6) | 17.0 (2.5) | .08 |
| **MMSE** | 25.8 (3.4) | 29.2 (1.0) | <.001 |

**Supplementary Table 2. Group comparison between ADNI patient and NC groups.**

Continuous variables are shown as mean (standard deviation). For comparisons across the three groups, $\chi^2$ tests of association were used for discrete and ordinal variables and t-tests were used for continuous variables.

#### Creating ROIs of connected regions

We defined regions connected to RSC and IP by performing a voxel-wise regression analysis in SPM similar to a metabolic covariance analysis[7,8] within the UCSF control group described previously. FDG SUVR in the RSC or IP was used as the independent variable and FDG SUVR in each voxel as the dependent variable. We then determined the MNI coordinates of peak correlation with the seed region, which we determined to be [-4, -46, 28] for RSC (located in the left hemisphere) and [56, -54, 22] for IP (located in the right hemisphere). These coordinates were inputted into *neurosynth.org*, a publicly available database of resting fMRI data for 1000 normal controls, to download maps of functional connectivity with each region. Final ROIs were determined by binarizing each map at the 90th percentile and removing the cluster of auto-correlation as described in the main text and in Figure S1.

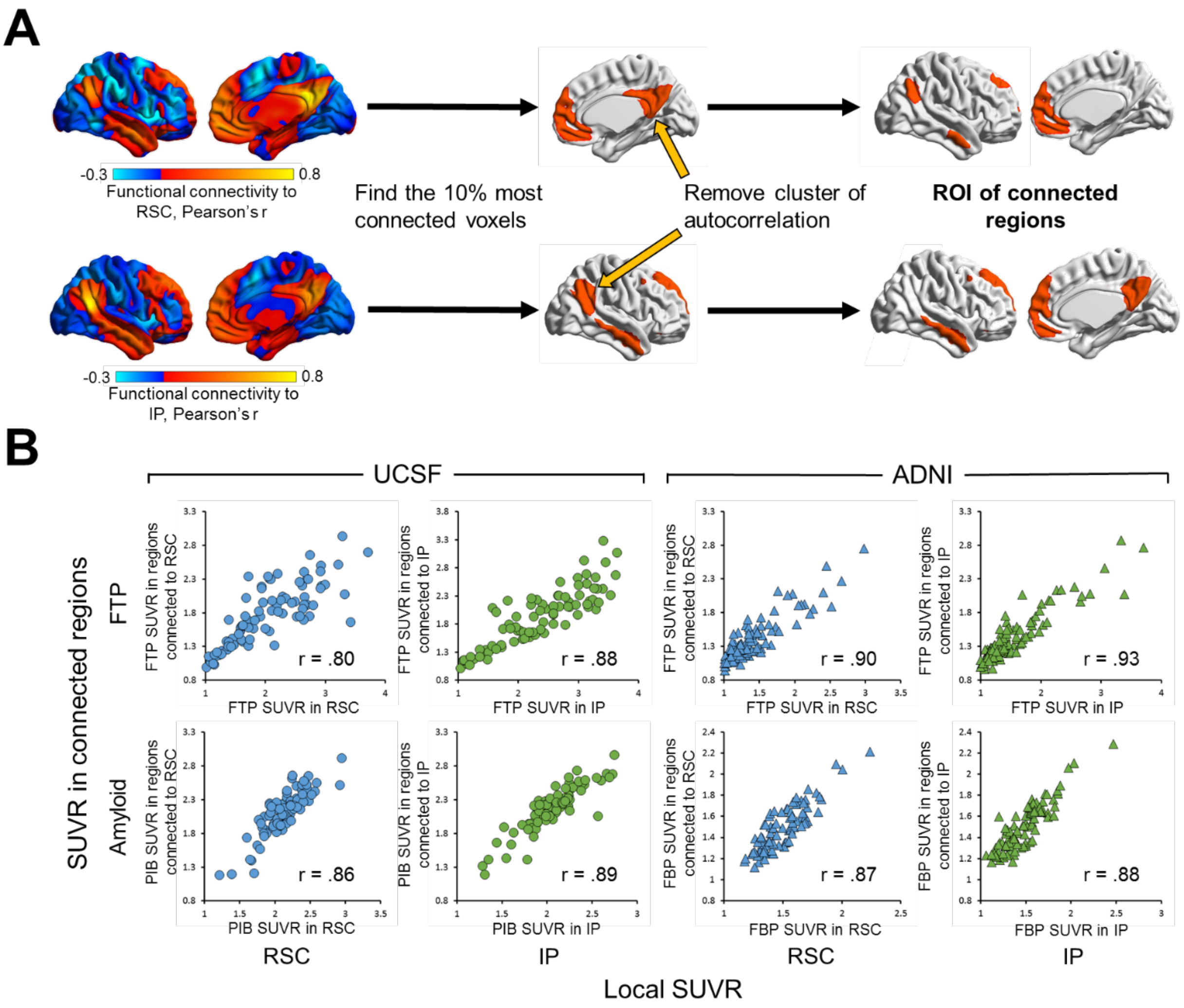


**Supplementary Figure 1. ROIs of connected regions.**

(**A**) Schematic to describe the creation of each ROI of connected regions (funcROI). (**B**) Scatterplots to describe the correlation between local FTP and amyloid-PET SUVR and the same metrics in connected regions, with FTP on the top and amyloid-PET on the bottom, UCSF on the left (circles) and ADNI on the right (triangles), and RSC associations in blue and IP in green.

#### Disease severity metric

The disease severity metric used aims to measure global impairment by combining functional (CDR-SB) and cognitive (MMSE) scores. Each score is obtained through different means and thus offers complementary information: CDR is obtained through a caregiver interview and MMSE is obtained through standardized cognitive tests with the patient. We first z-scored each score within cohort, reversed the MMSE-derived z-scores (so that higher values represent more severe deficits for both scores) then averaged the two z-scores across individual patients. We validated this composite metric by demonstrating that it improves correlation with FDG-PET compared to each score individually (Figure S2).

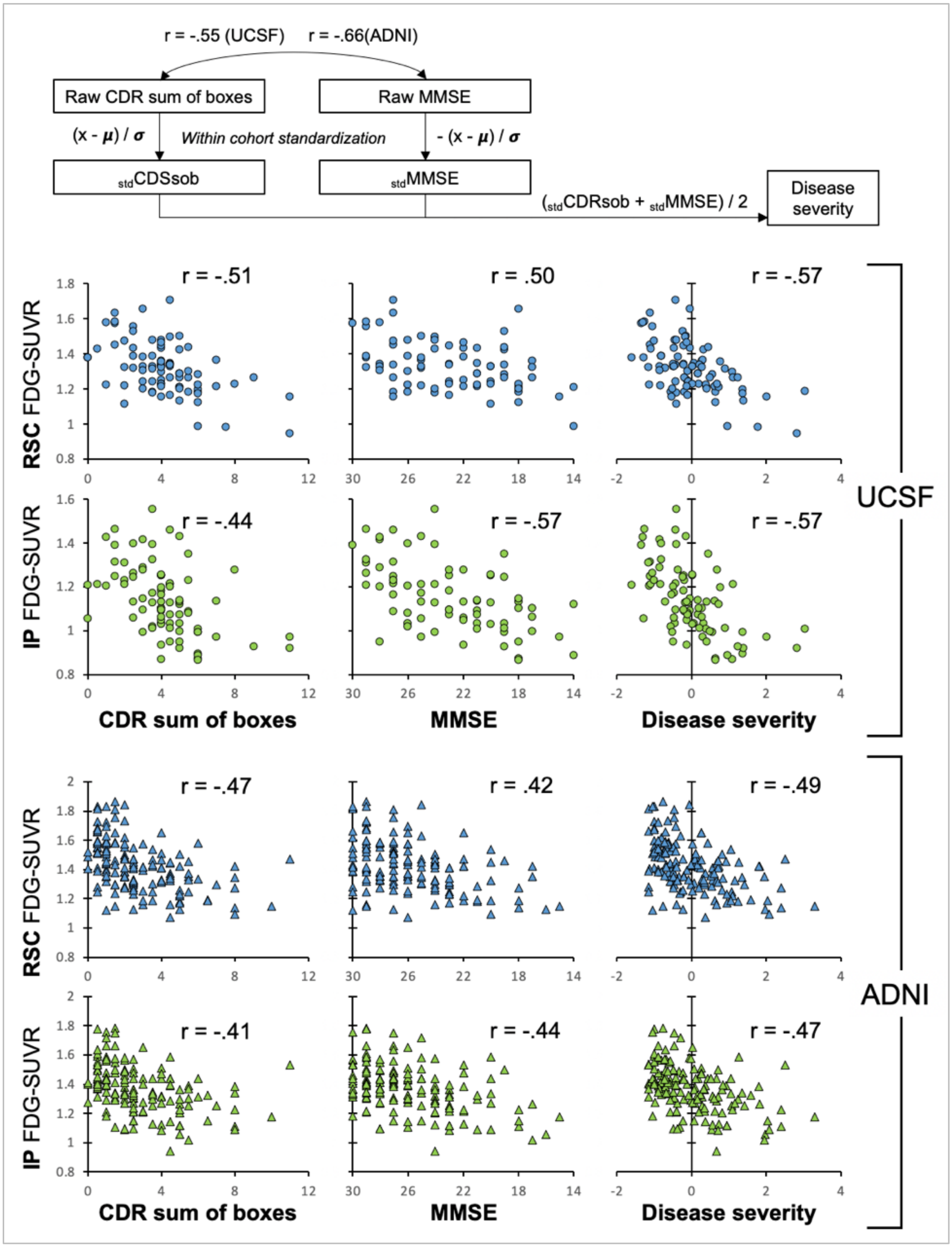


**Supplementary Figure 2. Disease severity metric derivation and validation.**

Combining MMSE and CDR-SB scores in describing disease severity improves correlations with FDG SUVR in primary regions of interest.

### Supplementary Results

#### Comparison of ADNI FBP and FBB subgroups

| | **ADNI FBP (n=87)** | **ADNI FBB (n=60)** | **Comparison** | |
|---|---|---|---|---|
| | | | Effect size | p |
| **Age** | 76.3 (7.9) | 74.5 (7.4) | d = 0.32 | .06 |
| **Sex (% F)** | 48% | 42% | V = 0.07 | .43 |
| **Education** | 15.7 (2.6) | 16.2 (2.5) | d = -0.18 | .25 |
| **CDR-Global (% ≥1)** | 24% | 17% | V = 0.09 | .28 |
| **CDR-SB** | 2.82 (2.3) | 2.64 (1.8) | d = 0.08 | .63 |
| **MMSE** | 25.6 (3.6) | 26.2 (3.0) | d = -0.19 | .25 |
| ***APOE* ε4 carrier (%, missing N)** | 60%, 9 | 80%, 14 | V = 0.21 | .02 |
| **Centiloid quantification** | 89 (41) | 82 (34) | d = 0.18 | .29 |

**Supplementary Table 3. ADNI subgroup demographic summary and comparison.**

Continuous variables are shown as mean (standard deviation). For comparisons between subgroups, $\chi^2$ tests of association were used for discrete variables (Cramer's V as effect size), Mann-Whitney tests were used for ordinal variables (Cohen's d as effect size) and t-tests were used for continuous variables (Cohen's d as effect size). CDR = Clinical Dementia Rating, CDR-SB = Sum of Boxes score, MMSE = Mini Mental State Exam, FBP = $^{18}$F-Florbetapir, FBB = $^{18}$F=Florbetaben.

#### Spatial correlation analyses

The similarity between patterns seen across cohorts or imaging modalities was quantified by extracting values from all 252,753 cortical voxels from the group and modality average maps (see neurovault: neurovault.org/collections/QNGOIQGC/) and running a correlation as illustrated in Figure 2. Multiple regression models were also run to explain patterns of hypometabolism using all three modalities.

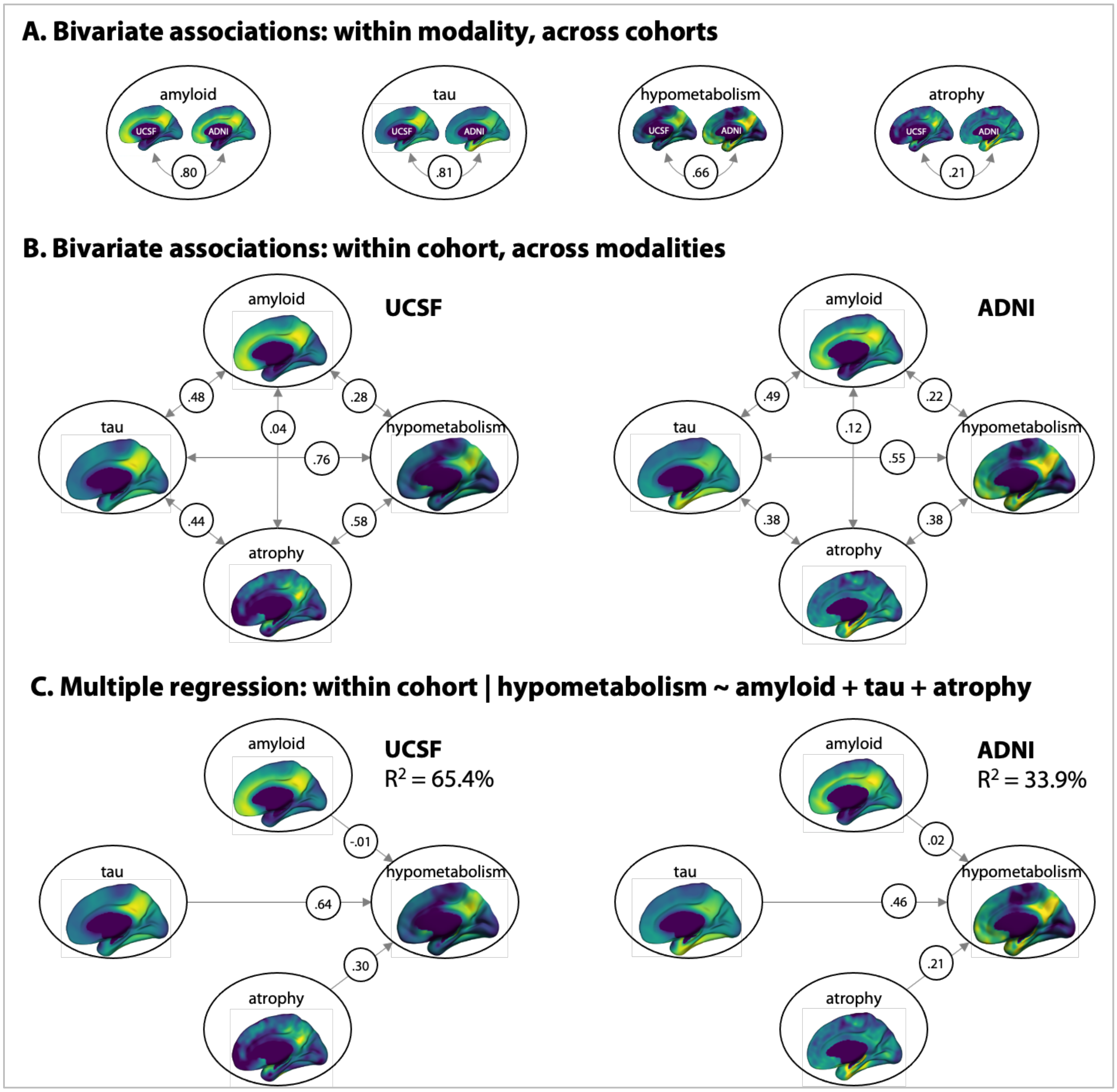


**Supplementary Figure 3. Spatial similarity between cortical imaging patterns.**

Circled values are standardized regression coefficients; in bivariate associations (**A** and **B**) they are equivalent to coefficient correlations, r. p values are not indicated as the extreme number of observations (252,753 voxels) make them irrelevant. Panel **A** includes data presented in Figure 2.

### PVC results

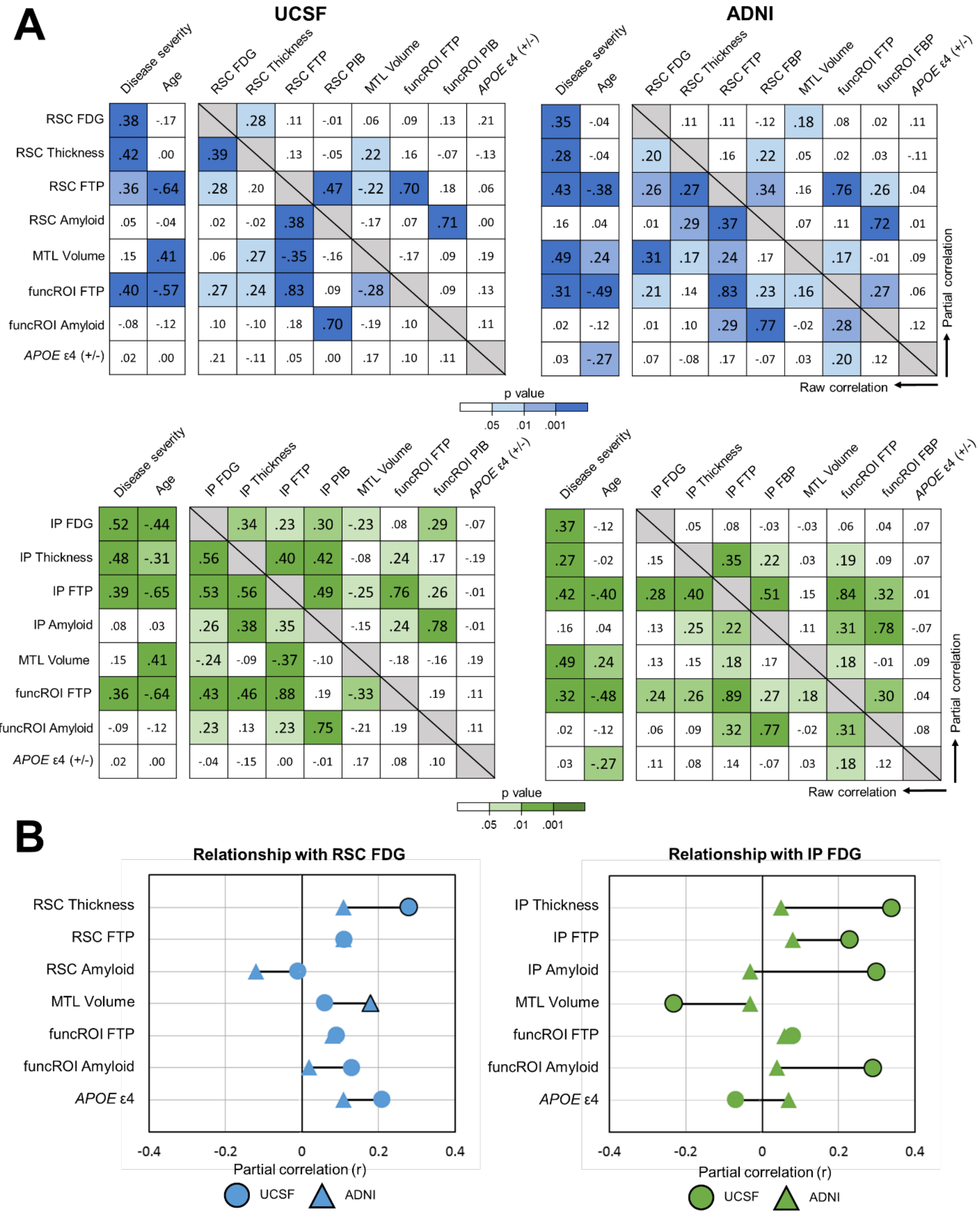


**Supplementary Figure 4. Bivariate and partial Pearson correlations using partial volume-corrected PET data.**

(**A**) Correlation matrices between all measures, with raw bivariate correlations in the bottom left portion and partial correlations controlling for age and disease severity in top right. Color saturation corresponds to p value. (**B**) Alternative presentation of partial correlations with FDG in RSC or IP with a direct visual comparison between ADNI (triangles) and UCSF (circle) cohorts. Shape border reflects significance ($p<.05$).

#### ADNI results using Florbetaben

The ADNI cohort consisted of 87 patients with FBP amyloid-PET and 60 with FBB amyloid-PET. The main amyloid-PET analyses were run within the FBP subgroup; we present the results for the FBB subgroup here.

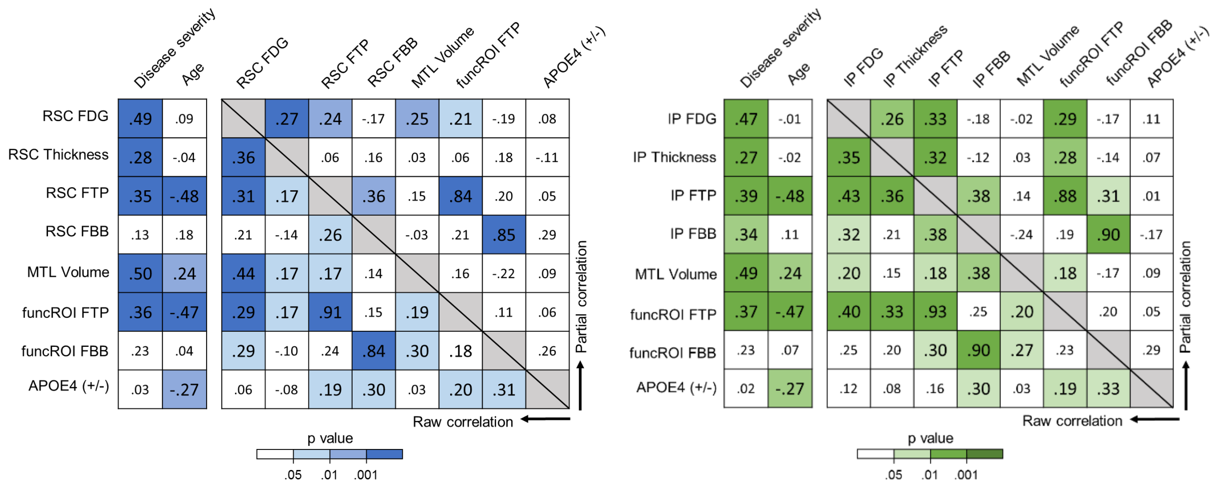


**Supplementary Figure 5. Correlation matrices with the ADNI FBB subgroup.**

Bivariate correlations in the bottom left portion of each matrix and partial correlations controlling for age and disease severity in top right. Left matrix is performed using the RSC ROI and right matrix using the IP ROI. Color saturation corresponds to p value. MTL volume was divided by total intracranial volume prior to analyses.

| **A. RSC FDG** | **Model 0** | | **Model 1** | | **Model 3** *(n=60)* | |
|---|---|---|---|---|---|---|
| **Predictors** | β | p | β | p | β | p |
| **Disease severity** | 0.48 | <.01 | 0.32 | <.01 | 0.32 | .01 |
| **Age** | 0.05 | .47 | 0.19 | .02 | 0.12 | .27 |
| **RSC thickness** | - | - | 0.24 | <.01 | 0.31 | <.01 |
| **RSC FTP** | - | - | 0.25 | <.05 | -0.08 | .74 |
| **FTP in connected regions** | - | - | - | - | 0.35 | .18 |
| **FBB in connected regions** | - | - | - | - | 0.20 | .06 |
| **$R^2$ ($\Delta R^2$)** | 0.24 (Ref) | | 0.33 (0.09) | | 0.51 (0.15) | |
| **BIC ($\Delta BIC_0$)** | -121 (ref) | | -130 (-9) | | -53 (0) | |

| **B. IP FDG** | **Model 0** | | **Model 1** | | **Model 3** *(n=60)* | |
|---|---|---|---|---|---|---|
| **Predictors** | β | p | β | p | β | p |
| **Disease severity** | 0.47 | <.01 | 0.29 | <.01 | 0.31 | .01 |
| **Age** | -0.05 | .48 | 0.12 | .17 | 0.11 | .35 |
| **IP thickness** | - | - | 0.16 | .04 | 0.17 | .16 |
| **IP FTP** | - | - | 0.32 | <.01 | 0.42 | .09 |
| **FTP in connected regions** | - | - | - | - | -0.02 | .95 |
| **FBB in connected regions** | - | - | - | - | 0.02 | .87 |
| **$R^2$ ($\Delta R^2$)** | 0.22 (Ref) | | 0.33 (0.11) | | 0.50 (0.19) | |
| **BIC ($\Delta BIC_0$)** | -134 (ref) | | -146 (-12) | | -58 (-2) | |

**Supplementary Table 4. Linear regression results with the ADNI FBB subgroup.**

Model 3 is performed in the FBB subgroup; whole-group Models 0-1 are included for comparison. Analyses are run within the ADNI cohort and within region, where FDG in either RSC (A) or IP (B) is the singular dependent variable. Local cortical thickness and FTP are included in all hypothesis-testing models due to their robust associations with FDG in previous correlation analyses. Model 3 is run within a smaller sample (n=60), and the reference levels for $R^2$ and BIC are modified accordingly. β = standardized estimate; BIC = Bayesian Information Criteria.
